# Effect of manual and digital contact tracing on COVID-19 outbreaks: a study on empirical contact data

**DOI:** 10.1101/2020.07.24.20159947

**Authors:** A. Barrat, C. Cattuto, M. Kivelä, S. Lehmann, J. Saramäki

## Abstract

In the fight against the COVID-19 pandemic, lockdowns have succeeded in limiting contagions in many countries, at however heavy societal costs: more targeted non-pharmaceutical interventions are desirable to contain or mitigate potential resurgences. Contact tracing, by identifying and quarantining people who have been in prolonged contact with an infectious individual, has the potential to stop the spread where and when it occurs, with thus limited impact. The limitations of manual contact tracing (MCT), due to delays and imperfect recall of contacts, might be compensated by digital contact tracing (DCT) based on smartphone apps, whose impact however depends on the app adoption. To assess the efficiency of such interventions in realistic settings, we use here datasets describing contacts between individuals in several contexts, with high spatial and temporal resolution, to feed numerical simulations of a compartmental model for COVID-19. We find that the obtained reduction of epidemic size has a robust behavior: this benefit is linear in the fraction of contacts recalled during MCT, and quadratic in the app adoption, with no threshold effect. The combination of tracing strategies can yield important benefits, and the cost (number of quarantines) vs. benefit curve has a typical parabolic shape, independent on the type of tracing, with a high benefit and low cost if app adoption and MCT efficiency are high enough. Our numerical results are qualitatively confirmed by analytical results on simplified models. These results may inform the inclusion of MCT and DCT within COVID-19 response plans.

## 1 Introduction

The COVID-19 epidemic was declared a pandemic by the WHO on March 11, 2020. At mid-July 2020, COVID-19 has reached at least 14 million confirmed cases worldwide and caused at least 600,000 deaths.^1^ Even if the situation appears under control in a number of countries and the first wave has been largely receding in many others, the epidemic is still accelerating in other parts of the world and its burden on healthcare systems is far from over.

Faced with an exponentially growing number of cases and given the absence of effective pharmaceutical treatment and lack of vaccine, governments have first had to rely on broad, nationwide measures aimed at reducing the mobility and number of contacts between individuals, starting with school closures and eventually lock-downs of whole countries.^2–7^ These non-pharmaceutical interventions (NPIs) have succeeded in limiting contagion^8^ and they have been gradually lifted in many cases. General measures to reduce transmission risk remain in place, such as enhanced hand hygiene, mandatory mask wearing in a number of contexts like public transportation and shops, and limits to the size of social gatherings.

These measures are beneficial and very useful instruments to avoid a resurgence and a second wave of the COVID-19 epidemic.^9^ They are also much easier to enforce than regional or country-wide lock-downs, and they do not carry their huge economic costs. However, such general measures are still indiscriminate: they impact infected and non-infected individuals equally. Targeted measures, on the other hand, focus on stopping the spread where and when it occurs. Many envisioned targeted interventions rely on the isolation of infectious individuals, which prevents the infected individual from further spreading the disease. This can be fully effective only if all infectious individuals are identified *before* they infect others (e.g. through symptoms or testing). However, in the case of COVID-19 a large fraction of transmissions occurs before symptom onset.^10–14^ Furthermore, many infected people only develop mild symptoms or no symptoms and are therefore typically not tested, despite being able to transmit the disease.^13,13,15^ A natural way to cast a wider net around an identified case is the so-called *contact tracing* process, which endeavors to identify people who have been in long enough contact with an infectious individual. The rationale is that those individuals, even in the absence of symptoms, might be infectious as well and that quarantining them and monitoring their health can both protect them and block further transmission.

Contact tracing is traditionally carried out via interviews of identified cases, followed by phone calls to the identified contacts to warn them and to ask them to go into quarantine.^16–18^ Such ‘manual’ contact tracing is labour-intensive; it can be slow, and it critically relies on the ability of individuals to remember and identify their contacts. While this is easy in some contexts, such as households, our actual ability to recall and identify close-range proximity contacts is known to be limited. Retrospective surveys have found that brief contacts have a lower probability of being recalled, and that contact durations are overestimated.^19–21^ Moreover, in important contexts such as public transportation, shops, or elevators, we often find ourselves in the proximity of unknown persons. In such cases, digital proxies for close-range proximity are currently viewed as a complementary and scalable approach, known as *digital contact tracing*, that could overcome the above limitations.^22–24^

The broad adoption of smartphones provides an opportunity to develop apps that use short-range device-to-device communication between phones to sense the proximity of their owners. The exchange of low-power Bluetooth packets between smartphones can indeed be used to detect proximity relations between persons whose phones are running the app: whenever a person who has installed a contact tracing app is diagnosed as infectious, a warning can be sent to all app users s/he has been in close proximity with during the previous few days.^22,25–27^ Different technical architectures for digital contact tracing have been proposed, including so-called “decentralized” solutions^25^ that by design minimize the amount of information that is collected and shared. These approaches have informed the design of the *Exposure Notification*^28^ mechanism by Apple and Google, who now jointly provide an Application Programming Interface (API) that national apps can use to carry out digital contact tracing. Exposed individuals identified via digital contact tracing are asked to contact the responsible health authorities and to quarantine themselves. Using a contact tracing app does not rely on individuals recalling or naming their contacts nor does it require knowledge of their identity. However, the efficiency of contact tracing apps is obviously limited by their numbers of users, since both the infectious person and their contacts need to have installed the app.

Some studies^27,29^ have proposed to further extend the reach of digital contact tracing by using information on individuals situated two hops away from a confirmed case, along the digitally-sensed proximity network (so-called “recursive” contact tracing). This might however have privacy implications, and it is important to note that the success of app-based contact tracing depends on a complex interplay of proximity tracing technology, citizens’ trust and adoption,^30^ effectiveness of the exposure notification strategy, and good integration with traditional contact tracing and with other public health capabilities and processes.

Several theoretical studies have investigated the potential effectiveness of proximity tracing apps in the fight against COVID-19.^17,18,22–24,27,31–33^ The studies that have most influenced the public discussion around contact tracing apps are based on a simplified, macro-level mathematical model that assumes homogeneous mixing of the population.^22,34^ However, real social and contact networks are highly non-homogeneous and exhibit many non-trivial structural and temporal features, including heterogeneities^20,35–37^ known to be highly relevant for epidemic spreading.^38–43^ While some studies have considered artificial static network structures to study at a theoretical level the effect of heterogeneous structures,^29,44^ other approaches have used large-scale agent-based models^32,45^ or GPS data^24^ to recreate synthetic daily aggregated networks of contacts and to evaluate the effectiveness of app-based contact tracing.

Using high-quality empirical data on close-range proximity between individuals is thus a crucial next step to improve the grounding of the discussion on the efficiency of targeted app-based interventions. State-of-the-art datasets of this kind were collected and made public over the years by several research efforts, in particular by two independent collaborations^37,46^ who have used proximity sensors and smartphones in specific communities (such as among university students) and in highly relevant social contexts such as workplaces, schools and hospitals. These empirical data are temporally resolved and exhibit all the complex structural and temporal complex features mentioned above.^20,35,36,47^ They represent thus an important benchmark for realistic simulations of epidemic processes and interventions. We note that these data have been recently used^33^ to derive parameters for the approach described in Ref.,^22^ and to determine – within that approach – which app-based contact tracing policies might be most effective.

Here, instead, we use such empirical high-resolution contact network data to inform a compartmental model for COVID-19^5^ and to simulate directly the targeted measures described above in realistic (albeit circumscribed) contexts. In particular, we study the combination of app-based and manual contact tracing, taking into account specific limitations of both approaches. We quantify the impact of interventions by measuring the reduction of the final size of the simulated epidemic. Moreover, we study the number of quarantines and the fraction of false positives, i.e., of quarantined individuals who were not infected. We find that the mere isolation of symptomatic cases is not sufficient to substantially reduce the epidemic size, while interventions guided by contact tracing can have a strong impact. We find that manual contact tracing (MCT), even when imperfect (i.e., limited recall of past contacts, delays in alerting contacts) yields a potentially strong reduction of epidemic size that grows linearly with the fraction of contacts correctly recalled. The effect of digital contact tracing, in turn, grows only quadratically with the fraction of app adopters, as expected from the constraint that both the case and the contact need to be running the app for a contact to be detected by their phones. Therefore, if digital contact tracing is used in isolation, a large fraction of adopters is required to obtain a large impact, although we report no threshold effects, and any degree of adoption yields a positive contribution. We moreover show that the overall qualitative behaviour of the epidemic size reduction can in fact be recovered in a simplified analytical model of propagation.

We also observe that manual and app-based contact tracing lead to similar numbers of quarantined individuals (that can be interpreted as the cost of the intervention) and fractions of false positives, for a given relative reduction of epidemic size (benefit of the intervention). In addition, such cost-benefit curve has a robust parabolic shape and exhibits a maximum: if contact tracing is effective enough, it might contain an outbreak early on, leading to few quarantine events, i.e., high benefit with low cost. We also provide analytical arguments to explain this robust parabolic shape. Overall, digital and manual contact tracing complement each other: digital contact tracing compensates for the inherent limitations in recall and scalability of manual contact tracing, but improvements in manual contact tracing efficiency can yield particularly strong effects.

Finally we show that, for fixed app adoption and manual contact tracing parameters, recursive contact tracing further reduces epidemic size, at the cost however of a higher fraction of false positives among quarantined individuals. In fact, even at given benefit (epidemic size reduction), the cost in terms of number of quarantines and false positives is higher for the recursive tracing than for the standard one. We conclude that the current best combination of targeted strategies is given by case isolation and manual contact tracing, whose limitations can be efficiently complemented by digital tracing. Improvement of the manual contact tracing efficiency should be prioritized, and app adoption should be as large as possible to reach a strong suppression of the spread, which can actually then occur at low cost in terms of quarantines.

## 2 Results

We model an outbreak of COVID-19 by means of a compartmental model^5^ in which individuals can be in a series of discrete states describing the unfolding of the disease (see Figure 1 and Methods for details and parameter values). Susceptible (healthy) individuals (S), upon contact with infectious ones, can contract the disease and first enter the exposed (non infectious) state (E) and then a pre-symptomatic infectious state (*I*_*p*_). Pre-symptomatic individuals can remain asymptomatic during the whole infectious phase (*I*_*a*_), with probability *p*_*a*_, or they can develop either mild or severe symptoms (*I*_*m*_ or *I*_*s*_), with respective probabilities *p*_*m*_ and 1 −*p*_*a*_ −*p*_*m*_. The infectious state leads to recovery R (or death) after a typical time.

**Figure 1:**
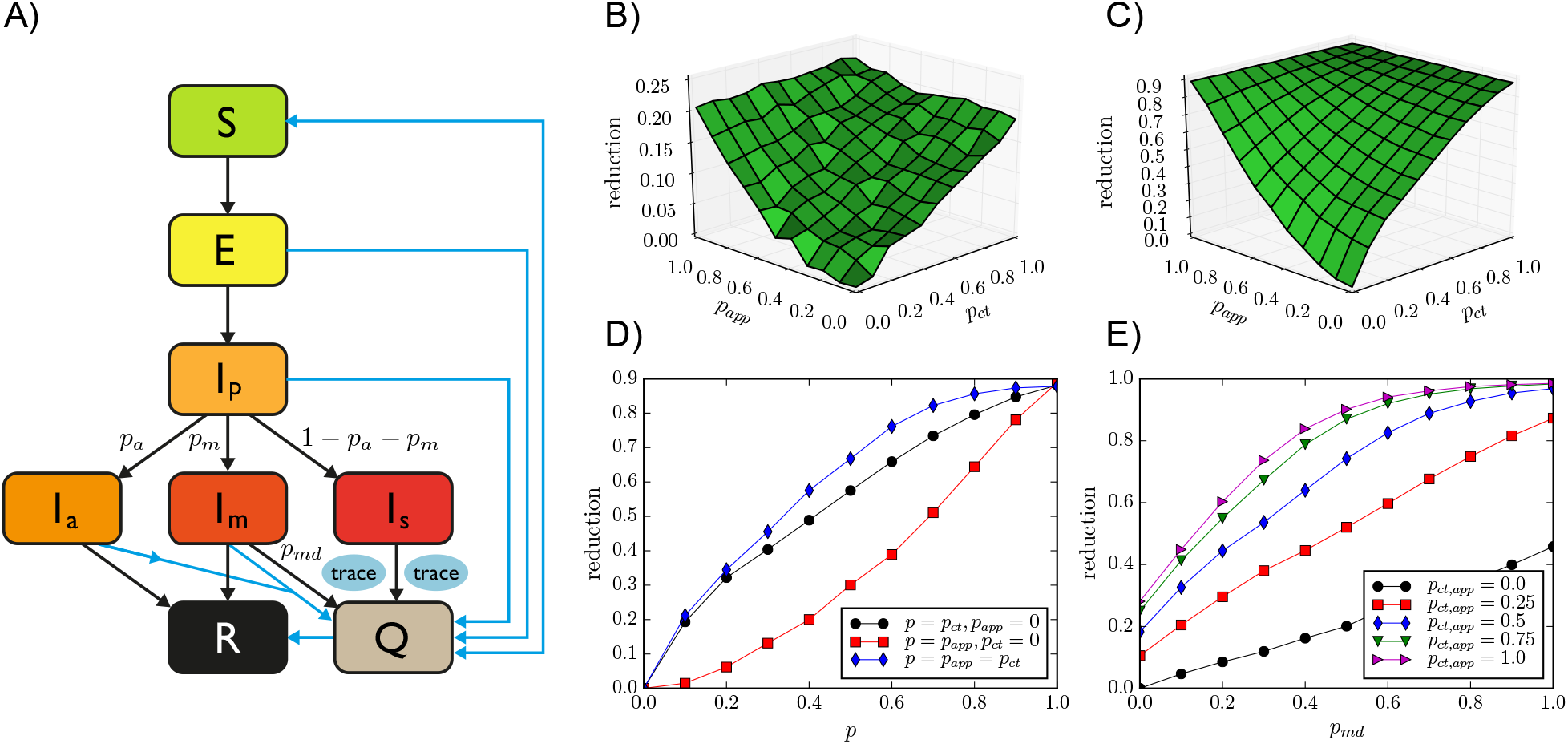
A) Schematic illustration of the investigated epidemic model. B) Relative reduction in the average epidemic size as a function of manual contact tracing probability *p*_*ct*_ and app adoption rate *p*_*app*_ for OD, with *θ*_*ct*_ = *θ*_*app*_ = 15 min. Here *p*_*md*_ = 0.5. C) The same for SD (threshold *θ*_*ct*_ = *θ*_*app*_ = 15 min). D) Same as B), for three slices: along the *p*_*ct*_ axis with *p*_*app*_ = 0, along the *p*_*app*_ axis with *p*_*ct*_ = 0, and along the diagonal *p*_*ct*_ = *p*_*app*_. E) Relative reduction as a function of the testing and diagnosis rate of mildly symptomatic *p*_*md*_, for the SD data with various values of *p_ct_* = *p_app_*.

We simulate the model on empirical data describing time-resolved, close-range proximity interactions of individuals in two relevant settings: a community of students (SD, in the following), and an office (OD, in the following). The SD dataset describes the network of physical proximity among a population of more than 700 university students over a period of four weeks. The dataset was collected as part of the larger Copenhagen Networks Study,^37,48^ in which each participant was equipped with a smartphone and required to install a data collection software. These devices were set to be permanently discoverable by Bluetooth and scanned for nearby Bluetooth devices at five minute intervals. Proximity was assessed by measuring the Received Signal Strength (RSSI) of Bluetooth signals from nearby devices: a high RSSI means that the two devices are physically close, a low measure indicates that devices are further apart or that there are obstacles in between.^49^ The OD dataset was collected in an office workplace staffed with about 200 people that agreed to wear for several days wearable proximity sensors while in the workplace;^46,50^ these sensors used low-power radio communication as a proxy for the close-range proximity of individuals wearing the devices.^35^ Both the SD and OD datasets represent close-range proximity interactions as a temporally-ordered sequence of contact networks, where nodes are individuals and an edge between nodes indicates a close-range proximity relation. For both datasets, the spatial resolution is of the order of 1-2 meters, making these datasets suitable for modeling at-risk contacts for droplet-transmitted pathogens such as SARS-CoV-2.

We run simulations starting from one initially infectious individual chosen at random, letting the spread evolve until no infectious individuals are present in the population. We first calibrate the parameters of the model to obtain a reproduction number *R*_0_ *≈* 3 in the absence of any containment measures.^5,51^ We also considered simulations with lower *R*_0_ values, corresponding, e.g., to re-emerging outbreaks during which a number of public health measures such as enhanced hygiene and mask-wearing are enforced to limit the spread. As a baseline intervention, used as a reference to assess the relative improvement of contact tracing, we then consider a simple strategy that does not rely on any contact information: isolation (Q) of all cases with severe symptoms as well as isolation of a fraction *p*_*md*_ of the mildly symptomatic cases, until they recover. For *p*_*md*_ = 50%, this intervention yields a relative decrease of the average epidemic size of, respectively, 14% for the OD data and 25% for the SD data.

We then simulate manual contact tracing (MCT) for all detected infectious cases (i.e., all severe cases *I*_*s*_ and a fraction *p*_*md*_ of the mild cases *I*_*m*_): for each case, an interview is assumed to be performed to identify all individuals who have been in contact with the case for an interval longer than *θ*_*ct*_ over the 48 hours before detection. To take into account the limitations of manual contact tracing, we assume that only a fraction *p*_*ct*_ of the contacts are identified, and that a delay of 2 days takes place between the detection of a case and the quarantining of a recalled contact. In addition to MCT, we consider the possibility of digital contact tracing (DCT): if the identified case has a smartphone with a contact tracing app installed, a warning is sent to other app users who have been in proximity (as assessed by the contact data) with the case for an interval longer than *θ*_*app*_ over the 48 hours prior to detection). Since we assume that all such contacts are identified by the app, the key parameter for DCT is the fraction *p*_*app*_ of the population who has adopted the app. While an ideal app would lead to an instantaneous quarantining of the warned individuals, we also consider potential delays due to possible app limitations (e.g., a limited number of exposure queries per day by the app).

The reduction of the average final epidemic size obtained by MCT and DCT, with respect to the situation of only case isolation, depends on the contact tracing parameters and efficiency. For simplicity we assume *θ*_*ct*_ = *θ*_*app*_ as these values are fixed by public health guidelines, and we vary the fraction *p*_*ct*_ of contacts identified by MCT and the fraction of app adopters *p*_*app*_ in the population. Note that, even if *p*_*app*_ in the general population is limited by the smartphone penetration, we consider here specific contexts where this penetration could be as high as 100% and we therefore explore the whole range of *p*_*app*_ values between 0 and 1.

Figure 1 shows how the reduction in epidemic size depends on these parameters. In particular, the effect is proportional to *p*_*ct*_ in the absence of digital contact tracing, while it grows quadratically with the fraction of app adopters in the absence of manual contact tracing. It is thus rather small at low app adoption and, in particular, improving MCT leads to stronger effects. However, even in the hypothetical scenario of perfect MCT, DCT still improves the results thanks to the shorter delay between detection and quarantine made possible by DCT, and the combination of DCT and MCT makes it possible to reach strong epidemic suppression effects even at intermediate parameter values.

We note that the qualitative behavior of the epidemic size reduction is similar for both datasets, despite the differences in contexts and data collection methods. In the SI, we show that this behavior is robust for other datasets and other parameter values.

Figure 1, moreover, illustrates the crucial role of detecting cases with mild symptoms. Even in the absence of tracing (*p*_*app*_ = *p*_*ct*_ = 0), increasing the probability of case detection increases linearly the reduction in the average epidemic size. This effect becomes even stronger when contact tracing is implemented and the combination of a large probability of detection and of contact tracing can make a strong suppression of the epidemic reachable. Finally, delays in the warning and quarantining of individuals using the app have only a very limited effect (not shown).

Contact tracing brings clear benefits as explored above, but has also a cost that can be quantified by the number of quarantine events in the population (note that a given individual might potentially be quarantined more than once). Moreover, a fraction of these quarantines are in fact unnecessary (“false positives”), as some of the quarantined individuals are, in fact, healthy. Figure 2 explores this issue, showing that the number of quarantine events, i.e., the cost of the intervention, initially grows with *p*_*app*_ and *p*_*ct*_, as expected. However, in some cases, this cost goes through a maximum and decreases for large enough DCT app adoption or MCT efficiency. This corresponds to the fact that a very efficient contact tracing procedure can actually stop an outbreak early on, so that the epidemic does not spread and few quarantines are necessary. In fact, Figure 2 shows cost-benefit curves of the contact tracing interventions, at fixed tracing parameters, i.e., the normalized number of quarantine events as a function of the reduction in average epidemic size. These curves all exhibit a typical parabolic shape, with a cost first increasing as a function of the benefit, reaching a maximum and then decreasing. Note that each curve puts together results obtained either by MCT, by DCT or by a combination of both: in other words, the cost depends on the benefit and not on the way (MCT or DCT) this epidemic size reduction was obtained. We also note on the other hand that the cost depends on the threshold *θ*_*ct*_ = *θ*_*app*_ used to define at-risk contacts in the contact tracing procedure: lower thresholds make it possible to obtain higher benefit at large adoption or MCT efficiency (large *p*_*ct*_), but the maximal value of the cost, reached at intermediate adoption or MCT efficiency, is then larger. Finally, increased delays in contact tracing leads to slightly increased costs, while very large values of case detection probability *p*_*md*_ allow to obtain both higher benefits and lower costs.

**Figure 2:**
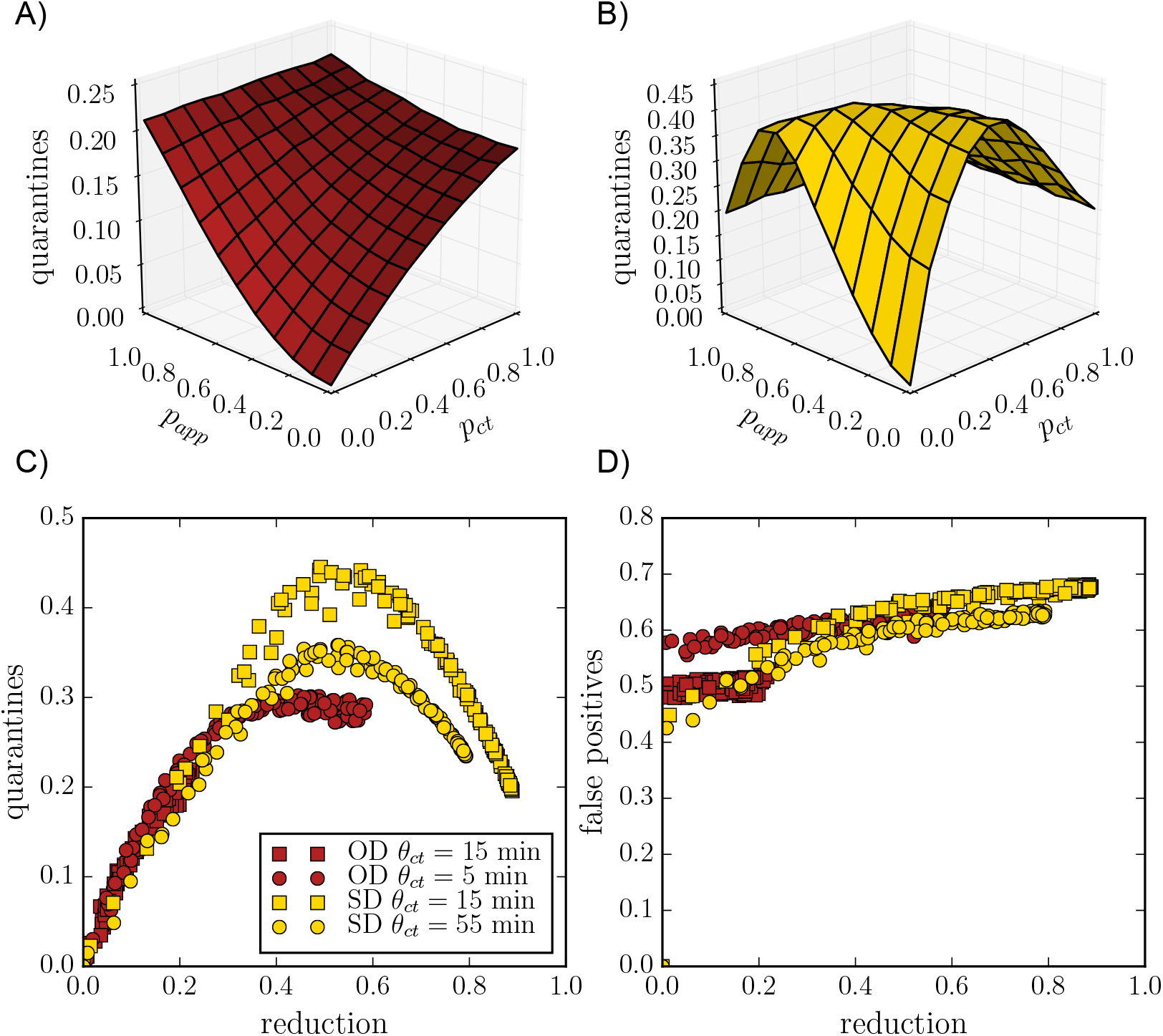
Number of quarantine events normalized by the population size, as a function of *p*_*app*_ and *p*_*ct*_, for A) the OD data and B) SD data. C) Number of quarantine events normalized by the population size as a function of epidemic size reduction, for the OD data (red squares; red circles for lower tracing threshold *θ*_*app*_ = *θ*_*ct*_ = 5 min), and SD data (yellow squares; yellow circles for higher threshold *θ*_*app*_ = *θ*_*ct*_ = 55 min)). D) Ratio of false positives to quarantines, *i*.*e*., the fraction of those quarantined but not ill, for the same data sets as in C).

Figure 2 shows also the behavior of the fraction of false positives. While this fraction only depends slightly on the app adoption and MCT efficiency, it increases slightly overall as the epidemic reduction increases. The fraction of false positives also increases if the delay between case detection and contact quarantining increases, and decreases if the detection probability of mild cases *p*_*md*_ increases.

While our results are obtained with simulations of a realistic compartmental model performed on state-of-the-art datasets describing human interactions, which include all the inherent complex features of such interactions, we can actually obtain a very similar phenomenology using analytical arguments on simplified models. To this end, we consider a Susceptible-Infectious-Recovered (SIR) model on a static random network. Using a well-known mapping of SIR models to percolation problems,^52^ it is indeed possible to obtain an equation for the final epidemic size of such a process. In the Supplementary Information we show that it is possible to extend the percolation arguments to introduce both digital and manual contact tracing, and we show in Fig. 3 that the resulting shape of the epidemic size reduction as a function of the tracing parameters *p*_*ct*_ and *p*_*app*_ closely reflects the phenomenology observed in our numerical simulations.

**Figure 3:**
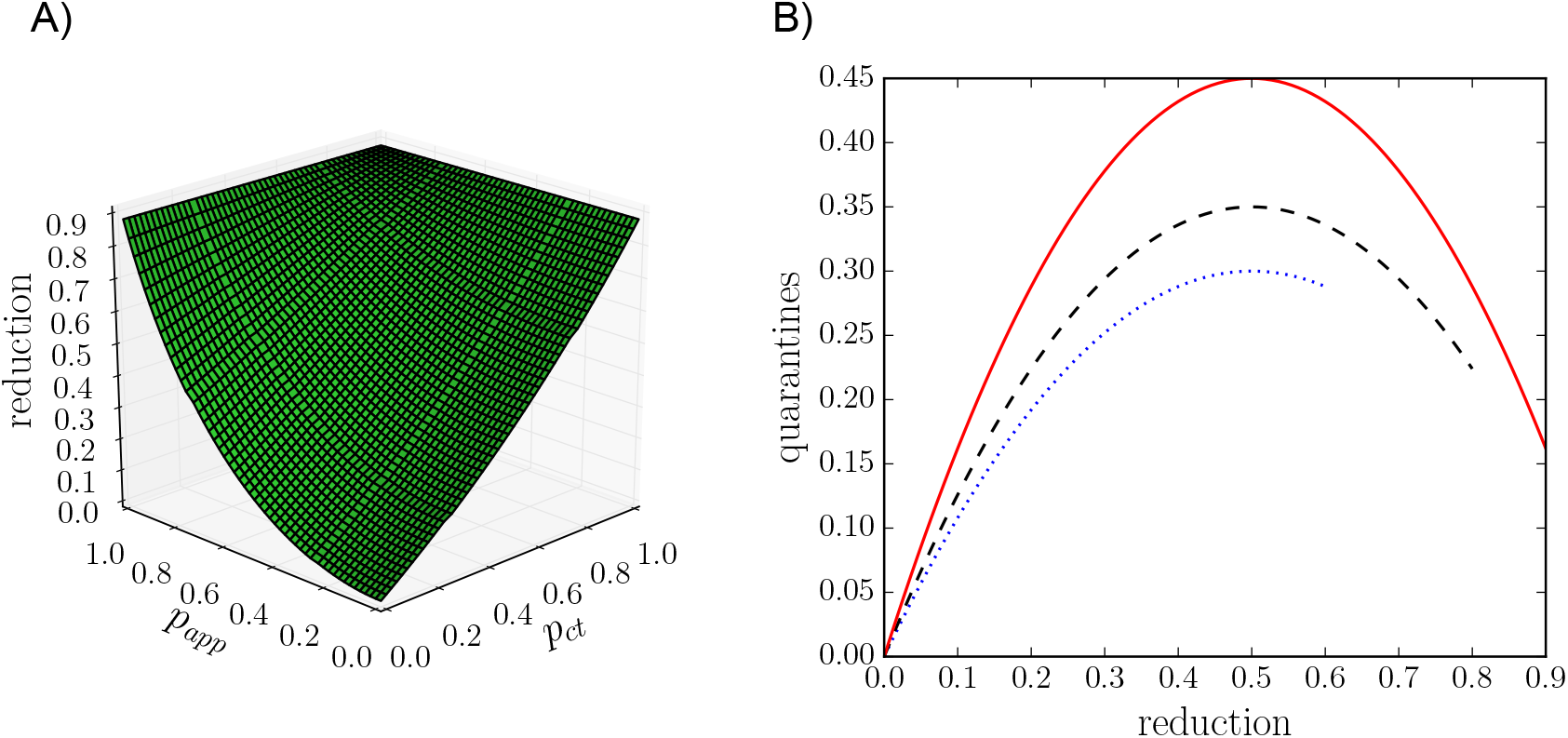
Results reproduced with stylised theoretical models. A) Reduction in final epidemic sizes as a function of manual contact tracing probability *p*_*ct*_ and app adoption rate *p*_*app*_. For easier comparison with the simulation results we have matched the parameters such that the epidemic sizes at *p*_*app*_ = *p*_*ct*_ = 0 and *p*_*app*_ = *p*_*ct*_ = 1 are the same as for the SD data shown in Fig. 1C. The overall shape of the surface is very similar, with manual contact tracing leading to almost linear reduction while the app adoption leads to a convex shape. B) Number of quarantine events normalized by the population size as a function of epidemic size reduction, obtained by the simple mathematical argument explained in the main text. The height of the curve depends on the parameters and the end of the curve is determined by the maximum reduction in epidemic size. For easier comparison with Fig. 2C we matched the heights and the maximum reductions to those of the simulation results.

Moreover, the parabolic shape of the cost-benefit curves shown in Fig. 2 can also be explained by a simple mathematical argument involving two competing effects (see SI for details). We denote the epidemic size reduction due to contact tracing as *r* = 1 − *I*_*r*_*/I*_0_, where *I*_0_ is the epidemic size without quarantine measures. First, as a mean-field assumption, we express the number of quarantines *q* as simply the product of the average number of contacts of a person during the infectious period *k*, the total number of infected people *I*_*r*_, and the quarantine probability *p*_*q*_, i.e., the probability that any contact with an infectious leads to a quarantine: *q* = *p*_*q*_*kI*_*r*_. Quarantines however yield a decrease in the epidemic size so that the number of infectious people is itself a decreasing function of the quarantining probability: *I*_*r*_ = *f* (*q*_*r*_)*I*_0_. Figure 1 shows that the function *f* is roughly linear in first approximation. Combining these two competing factors and solving for *q* as a function of *r* leads to the quarantine curve *q*(*r*) *∝ r* − *r*^2^, which yields a downwards opening parabola very similar to Fig. 2, as shown in Fig. 3. More details and examples are provided in the Supplementary Information.

We finally show – in Figure 4 – the impact of a recursive digital contact tracing. At fixed *θ*_*app*_ and app adoption, the obtained reduction of the epidemic size is larger than with the normal app. The difference relative to the usual DCT is however quadratic in *p*_*app*_, meaning that this effect remains small unless app adoption is very high. Moreover, the resulting number of quarantine events is larger and depends more on the other parameters of the contact tracing. In fact, Figure 4 shows that, at a given value of the epidemic size reduction, the number of quarantines is typically larger for the recursive DCT than for the usual DCT, and that the fraction of false positives, i.e., of unnecessary quarantines, becomes larger as well. In other words, the cost to obtain a given benefit is higher.

**Figure 4:**
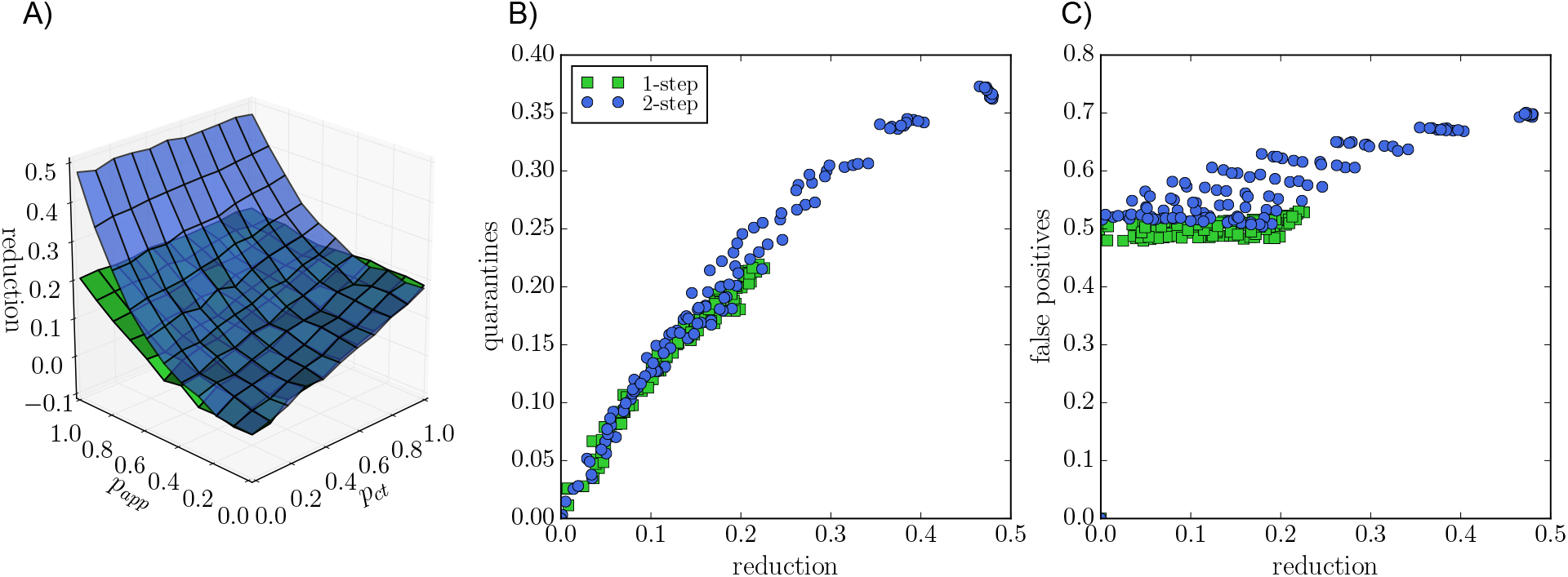
Recursive contact tracing versus single-step contact tracing. A) Blue surface: epidemic size reduction as a function of manual contact tracing probability *p*_*ct*_ and app adoption rate *p*_*app*_ for recursive tracing (blue) and single-step tracing (green) for reference, for the OD data set. *θ*_*ct*_ = *θ*_*app*_ = 15 min. B) Normalized number of quarantine events versus epidemic size reduction for recursive (“2-step”, blue) and single-step tracing (green). C) Proportion of false positives for the same cases.

## 3 Discussion

In the fight against the COVID-19 pandemic, non-pharmaceutical interventions play a crucial role. Moreover, in order to limit societal costs, targeted measures such as isolation of cases, contact tracing and quarantining of contacts are considered as essential in containing potential re-emergence of outbreaks. As traditional manual contact tracing is labour extensive and limited by people’s ability to correctly remember contacts, app-based digital contact tracing is seen as a potentially useful complement and has been deployed in several countries. Its actual efficiency, however, has been debated, in particular with respect to the level of adoption needed for it to make a difference.

In this study, we have provided a new quantification of the efficiency of contact tracing by leveraging state-of-the-art datasets describing contacts between individuals in different settings, namely among office workers and among students of a university. Indeed, most previous works have considered either homogeneous mixing hypothesis or model networks of interactions, while real contacts are known to display a wealth of structural and temporal heterogeneities having an impact on epidemic propagation. The use of real contact data in simulations is therefore a crucial step in the evaluation of interventions. We have considered a compartmental model drawn from the most recent literature on the propagation of the SARS-CoV-2 pathogen, and simulated several mitigation measures, focusing on the reduction in the final average epidemic size in the population. The isolation of severe cases and of a fraction of mildly symptomatic cases, although beneficial, is not enough to contain the spread as pre-symptomatic individuals are known to be infectious, and as a fraction of infectious are moreover asymptomatic. We have then simulated contact tracing, both manual (MCT) and digital (DCT), and shown that both can yield a strong decrease in the final epidemic size. This effect depends on the efficiency of the manual tracing, quantified by the fraction of contacts recorded for each detected infectious individual, and on the app adoption in the case of digital tracing. We find that the reduction of epidemic size grows linearly with the efficiency of MCT but only quadratically with the app adoption, as both case and contact need to have the app installed for a contact to be detected. We have shown that, at a qualitative level, this overall behaviour can be recovered analytically in a simplified epidemic model. We note that, as our study focuses on specific contexts (offices and university), the app adoption could potentially reach 100% in the corresponding populations, leading in that case to a very strong suppression of the epidemic. We have also shown that the cost of the intervention, as quantified by the number of quarantines, initially grows as the MCT efficiency and app adoption increase, but can become smaller if these parameters are high enough so that the spread is very efficiently contained. The fact that the cost-benefit curves show a typical parabola shape that can be understood using simple arguments: for low efficiencies of contact tracing, the cost increases with the reduction of epidemic size, but if the efficiency becomes large enough, the strong suppression of the outbreak leads to fewer cases, thus fewer contacts and fewer quarantines. Slightly more formally, we have shown how a simple analytical argument recovers this parabolic shape of the cost-benefit curves.

We also note that the digital contact tracing we have simulated does not imply knowledge of the contact network of infectious individuals, but simply that their contacts receive a warning and quarantine accordingly. This shows that it is possible to develop strongly privacy-preserving protocols^25^ that might reach large app adoption levels and thus yield a strong impact. Further, we have shown that recursive DCT yields an increased impact which, however, grows also only quadratically with app adoption. The added benefit remains thus small except at very high adoption, and comes at an increased cost in terms both of quarantine events and of false positives. Moreover, it is important to remark that recursing the CT process over contacts of an index case entails building an explicit representation of the 2-hop ego network of confirmed cases, exposing significantly more network information about those individuals than regular CT. This additional network information dramatically increases privacy risks, as it can be more readily used to match the contact graph around a given user to social network data from other sources (e.g., on-line social networks, mobile call networks, organizational networks, etc.), increasing the probability of re-identification. The resulting privacy concerns might lead to lower app adoption in the public, and thus potentially to an actual loss of efficiency of the digital contact tracing efforts.

Our study has a number of limitations which are important to make explicit. In particular, the data we use correspond to limited social environments (a university campus and a workplace) and we do not provide an overall general study that includes all possible multiple and differentiated contexts and their mutual interplay. These data moreover do not include interactions between the individuals participating in those studies and the general population. Nevertheless, these datasets correspond to the current state-of-the-art in terms of data describing human interactions, and the results show a robust qualitative behavior for data obtained in different contexts with different data collection infrastructures, both in terms of the epidemic size reduction and how the cost of the intervention depends on the manual contact tracing efficiency and on the app adoption.

Although the effect of DCT is only quadratic in the app adoption, we emphasize that there is no threshold effect: any increase in adoption leads to an improvement in the epidemic spread mitigation. This is also true for MCT, showing that the quality of interviews and any improvement in the ability to correctly find contacts of infectious cases brings an important benefit. Overall, DCT, which is also able to trace contacts between individuals who do not know each other, yields an interesting complement to MCT, and the combination of MCT and DCT is able to suppress outbreaks at limited cost if the app adoption and MCT efficiency are sufficiently high.

As the app adoption in the general population is necessarily limited by smartphone penetration, it is finally interesting to emphasize that this limitation does not necessarily hold in a specific context such as a workplace or a university. In fact, one could expect assortativity or group effects such that the app adoption is very high in such specific contexts or populations. This could make it possible to very rapidly suppress outbreaks in these specific populations, at minor costs in terms of quarantines. A full investigation of such assortativity effects is an interesting direction for future work, and could lead to policy indications on how to best focus testing and MCT resources towards populations where app adoption is expected to be lower.

## Data and Methods

### Data

We use state-of-the-art datasets describing contacts between individuals in different settings, with high spatial and temporal resolution.

- The SD dataset^37,48^ describes the interactions of 706 students, as registered by the exchange of Bluetooth signals between smartphones, for a period of one month. Each participant in the study was equipped with a Google Nexus 4 smartphone (used as primary phone) and required to install study-designed data collection software. For Bluetooth measurement, all devices in the experiment were configured to be permanently discoverable, and to scan for nearby Bluetooth devices at five minute intervals. The devices measured and recorded the Received Signal Strength (RSSI): a high RSSI means that the two devices are physically close, a low measure indicates that devices are further apart or that there are obstacles in between.^49^
- The OD dataset was collected by the SocioPatterns collaboration, using an infrastructure based on wearable sensors that exchange radio packets, detecting close proximity (≤ 1.5*m*) of individuals wearing the devices.^35^ The temporal resolution of the data is of 20 seconds. The data were collected among 232 individuals in offices during 2 weeks in 2015.^47^ If the spread simulated on the data lasts more than the dataset duration, we simply replicate it.^42,43^
- We show in addition in the Supplementary material results obtained using data collected among more than 300 students in a French High School during one week by the SocioPatterns collaboration, i.e., with the same infrastructure as the OD data.^20^

### Epidemic model

We consider the compartmental epidemic model described in Ref.,^5,6^ which was designed to describe the various stages of the COVID-19 disease.

Susceptible individuals (S) can become infected upon contact with infectious one, and enter then a latent phase (E) of duration *τ*_*e*_. They become then pre-symptomatic (*I*_*p*_) during *τ*_*p*_. Pre-symptomatic can then become either asymptomatic (*I*_*a*_), or develop mild or severe symptoms, entering respectively the compartments *I*_*m*_ or *I*_*s*_. This happens with probabilities *p*_*a*_, *p*_*m*_, *p*_*s*_ = 1−*p*_*a*_ −*p*_*s*_. Recovery occurs after a time 1*/µ*.

The probability per unit time of a susceptible to become infectious upon contact with an infectious with severe symptoms is *β*. Upon contact with a presymptomatic who will later on develop severe symptoms, the probability per unit time becomes *r*_*p*_*β*, and it is *r*_*β*_*β* upon contact with an asymptomatic or an infectious with mild symptoms, or with a presymptomatic who will then develop mild or no symptoms.

The parameter values are given in Table 1. The value of *β* is adjusted in order to fix the basic reproduction number to a desired value in the absence of interventions. For *R*_0_ = 3 this yields *β* = 1.37*e* − 3 for OD and *β* = 6.25*e* − 3 for SD.

**Table 1:** Parameters of the compartmental model

| Parameter | Value |
| --- | --- |
| $\tau_e$ | 3.7 days |
| $\tau_p$ | 1.5 days |
| $1/\mu$ | 2.3 days |
| $R_0$ | 3 |
| $p_a$ | 0.2 |
| $p_m$ | 0.72 |
| $r_\beta$ | 0.5 |
| $r_p$ | 1 |

### Interventions

We consider here a series of targeted interventions, i.e., that aim at preventing further transmissions from infected individuals.

#### Isolation

The first intervention consists simply in isolating infectious individuals once they are identified. This happens with all infected developing severe symptoms, as they will reach out to health services. Moreover, it can also happen for a fraction of the infected developing mild symptoms. The probability that an individual with mild symptoms is identified as infectious is *p*_*md*_. Moreover, we take into account that the reaction to symptoms is not instantaneous by introducing a delay *τ*_*to isol*_ between the appearance of symptoms and the isolation. Upon isolation, an infectious stops having contacts and becomes unable to transmit the disease.

Pre-symptomatics and asymptomatics cannot be identified and therefore are not isolated.

#### Manual contact tracing

When an infected individual (a “case”) is identified, s/he is interviewed by healthcare workers and asked to remember her/his contacts of the last *n*_*keep*_ days. The persons who have been in contact with the case for a cumulative time longer than *θ*_*ct*_ during these days are then warned and asked to quarantine for *τ*_*q*_ = 14 days.

The imperfection of manual contact tracing is taken into account by the following parameters:

- only a fraction *p*_*ct*_ of the contacts longer than *θ*_*ct*_ is recalled;
- only a fraction *p*_*c*_ of the contacts agree to quarantine; the others do not act;
- there is a delay *τ*_*ct*_ between the detection of the case and the quarantine of her/his identified contacts.

The health of quarantined individual is monitored. Therefore, if a quarantined person develops symptoms (even mild), s/he is interviewed to trace her/his recent contacts, with the same procedure as the initial case. The parameter values are given in Table 2.

**Table 2:** Parameters of the interventions. We also explore *p*_*c*_ = 0.6 and the whole range 0 ≤ *p*_*md*_ ≤ 1.

| Parameter | Value |
| --- | --- |
| $\tau_{to\ isol}$ | 0.5 day |
| $p_{md}$ | 0.5 |
| $\tau_q$ | 14 days |
| $n_{keep}$ | 2 days |
| $p_c$ | 1 |
| $\tau_{ct}$ | 2 days |

#### Digital contact tracing

We consider, either by itself or in addition to manual contact tracing, the possibility that individuals have installed a privacy preserving proximity tracing app. The app registers proximity events with other individuals equipped with the app.

Whenever an app-adopter is diagnosed as infected (the case), the anonymous random identifiers used by her/his app in the last *n*_*keep*_ days are uploaded to the central server (see^25^ for details).

All app-adopters regularly check the central server and their app compares the list of identifiers of infected individuals to the list of identifiers received in the previous days. If the app detects that the cumulated time in contact with (one or several) infected app-adopters in the last *n*_*keep*_ days exceeds *θ*_*app*_, it triggers a warning to contact health authorities and go into quarantine.

We assume that the transmission of information is instantaneous and that compliant individuals start immediately their quarantine. The parameters are the fraction of app-adopters in the population, *p*_*app*_, and the probability *p*_*c*_ that an individual receiving a warning goes into quarantine. Non-compliant individuals do not change their behaviour. If a quarantined app-adopter develops symptoms (even mild), the process is iterated, i.e., her/his identifiers are uploaded. The parameter values are given in Table 2.

#### Recursive contact tracing

We finally implement recursive (2-steps) warning for app-adopters: in this case, we assume that a protocol is in place so that app-adopters can compute the cumulative contact time during the previous *n*_*keep*_ both with app-adopting infected individuals and with app-adopting contacts of app-adopting infected individuals that have received a warning by the app. If this cumulative time exceeds *θ*_*app*_, it triggers a warning to contact health authorities and go into quarantine.

The parameters are the same as for the digital contact tracing: the fraction of app-adopters *p*_*app*_, and the probability *p*_*c*_ that an individual receiving a warning goes into quarantine. Non-compliant individuals do not change their behaviour.

### Quantification of the impact of interventions

For each scenario, defined by a given set of interventions with a given set of parameter values, we perform 10, 000 numerical simulations with a single, randomly chosen individual in the latent phase at the initial time. Simulations are run until no infected individuals are present in the population, i.e., all individuals are either susceptible or recovered.

In order to quantify the impact of the spread on the population, and the effect of the various interventions, we measure:

- the average final epidemic size, i.e., the fraction of individuals who are recovered at then end of the simulation;
- the fraction of population going through quarantine;
- the fraction of the quarantined individuals who were in fact susceptible (“false positives”).

## Data Availability

All data is publicly available

## Acknowledgements

This study was partially supported by the Lagrange Project of the ISI Foundation funded by CRT Foundation to CC. It was partially supported by the ANR project DATARE-DUX (ANR-19-CE46-0008-01) to AB.

